# Ethnicity and risk of death in patients hospitalised for COVID-19 infection in the UK: an observational cohort study in an urban catchment area

**DOI:** 10.1101/2020.05.05.20092296

**Authors:** E Sapey, S Gallier, C Mainey, P Nightingale, D McNulty, H Crothers, F Evison, K Reeves, D Pagano, A.K Denniston, K. Nirantharakumar, P Diggle, S Ball, on behalf of all clinicians and students at University Hospitals Birmingham NHS Foundation Trust

## Abstract

**Background:** Studies suggest that certain Black and Asian Minority Ethnic groups experience poorer outcomes from COVID-19 but these studies have not provided insight into potential reasons for this. We hypothesised that outcomes would be poorer for those of South Asian ethnicity hospitalised from a confirmed SARS-CoV-2 infection, once confounding factors, health seeking behaviours and community demographics were considered and that this might reflect a more aggressive disease course in these patients.

**Methods:** Patients with confirmed SARS-CoV-2 infection requiring admission to University Hospitals Birmingham NHS Foundation Trust(UHB) in Birmingham UK between 10^th^ March 2020-17^th^ April 2020 were included. Standardised Admission Ratio(SAR) and Standardised Mortality Ratio(SMR) were calculated using observed COVID-19 admissions/deaths and 2011 census data. Hazard Ratio (aHR) for mortality was estimated using Cox proportional hazard model adjusting and propensity score matching.

**Results:** All patients admitted to UHB with COVID-19 during the study period were included (2217 in total). Fifty-eight percent were male, 69.5% White and the majority (80.2%) had co-morbidities. Eighteen and a half percent were of South Asian ethnicity, and these patients were more likely to be younger, have no co-morbidities but twice the prevalence of diabetes than White patients. SAR and SMR suggested more admissions and deaths in South Asian patients than would be predicted and they were more likely to present with severe disease despite no delay in presentation since symptom onset. South Asian ethnicity was associated with an increased risk of death; both by Cox regression (Hazard Ratio 1.4 (95%CI 1.2–1.8) after adjusting for age, sex, deprivation and comorbidities and by propensity score matching, matching for the same factors but categorising ethnicity into South Asian or not (Hazard ratio 1.3 (1.0-1.6)).

**Conclusions:** Those of South Asian ethnicity appear at risk of worse COVID-19 outcomes, further studies need to establish the underlying mechanistic pathways.

## Introduction

Coronavirus disease 2019 (COVID-19) was identified in January 2020 (1) and given its designated name by the World Health Organisation in February, 2020(2). Initial reports from China, Italy and the USA focused on risk factors which predisposed individuals to severe manifestations of infection, including viral pneumonia and adult respiratory distress syndrome (ARDS) requiring critical care support and death, including age, male sex and co-morbidities (3–6).

A more recent report from the Centers for Disease Control and Prevention (CDC) in the USA described an early signal of non-Hispanic, Black people being disproportionally affected by COVID-19 hospitalisation(7). In the UK, the Intensive Care National Audit and Research Centre (ICNARC) reports(8, 9) described that a higher proportion of patients requiring critical care for COVID-19 were of Asian and Black ethnicity compared to pre-COVID historic data (2017 - 2019) for patients who required critical care for viral pneumonia. Since then two studies, one from Office for National Statistics analysing over 10,000 deaths in UK (10) and another examining around 5,000 deaths from primary care records(ll) concluded two to three fold high death rates in ethnic minority groups after accounting for important confounders. However, both studies reported mortality at a population level that could be a reflection of higher infection rates in these ethnic groups rather than a higher case fatality rate.

The West Midlands is experiencing a high incidence of COVID-19 associated hospitalisations. This is particularly marked in Birmingham which has a higher than average percentage of minority ethnic groups, with the 2011 census reporting the following percentages: South Asian 23.4%, Black 7.8%, Mixed ethnicity 4.1%, Others 1.8% and a lower than average White ethnic group at 63% (12).

University Hospitals Birmingham NHS Foundation Trust (UHB) is one of the largest NHS Trusts in England, providing direct acute services and specialist care across four hospital sites, including 2.2 million patient episodes per year, with 2750 beds and an expanded ITU capacity of up to 250 beds during the COVID-19 pandemic. UHB constitutes 4 acute hospital sites following organisational merger in 2018. At present the Queen Elizabeth Hospital Birmingham (QEHB) runs a fully electronic healthcare record (EHR) (PICS; Birmingham Systems-in place since 1999), while the other three sites currently run mixed electronic and paper healthcare records and a shared primary and secondary care record (Your Care Connected). UHB provides secondary care to a diverse population of 1.3 million in Birmingham and Solihull and provides a full range of tertiary services to the WM region.

It was hypothesised that South Asian ethnicity would form a risk factor for the most severe respiratory manifestations of COVID-19 infection, even once age, sex, medical conditions and social deprivation were taken into account and therefore:

a. Have more admissions than would be expected given the proportion of different ethnic groups within the Birmingham Trust catchment area (based on the last census data).
b. Have worse outcomes (death and/or admission to critical care) from hospitalised COVID-19 viral infection than White ethnic group, even once age, gender, deprivation and co-morbidities were accounted for.
c. Have more severe disease on presentation based on a severity score which could not explained by duration of symptoms, compared to non-South Asian patients.

The study had the following aims.

1. To identify all COVID-19 confirmed patients admitted to UHB hospital within a determined timeframe
2. To determine the expected and observed admission and death rate given the local population
3. To determine whether South Asian ethnicity was associated with poor outcomes following hospitalisation with confirmed COVID-19 infection, once potentially confounding factors were considered.
4. To explore if disease presentation was more severe in patients of South Asian ethnicity and if there was any evidence of a delayed presentation to hospital.

## Methods

This retrospective cohort study, using prospectively collected data was conducted in affiliation with PIONEER (, the HDR-UK Health Data Research Hub in acute care.

### Study population

All patients with a confirmed positive severe acute respiratory syndrome coronavirus-2 (SARS-CoV-2) swab result between 9:00am 10^th^ March 2020 and 16:00 17^th^ April 2020, and who were admitted to UHB at the time of or up to two weeks following their first positive SARS-CoV-2 swab test were included. COVID cases were confirmed following a nasopharyngeal and oropharyngeal swab in all cases(13) which were processed in accordance with NHS guidance within UHB NHS laboratories (14). Mortality and (in those alive) patient admission status (discharged and alive, continued admission and alive) were assessed on 12^th^ May 2020.

### Data Collection and variable definitions

Patient demographics and clinical data were collected from the EHR and from mandator datasets within the Trust. Clinician confirmed co-morbidities were available from the EHR, the depth of which was enhanced by access to a summary primary care record (Your Care Connected) and further enriched with diagnostic codes derived from previous hospital episodes. The EHR encodes diagnoses using NHS Digital SNOMED CT browser(15) alongside and mapped on to ICD-10 codes(16) allowing for the presentation and inclusion of historically entered ICD10 codes. Co-morbidities of interest were defined by those associated with poor outcomes from previous publications (17, 18) in order to determine the impact of multi-morbidity(18). The most common clusters of diagnostic categories are listed in Table 1. A simple count of co-morbidities was undertaken to determine the impact of multi-morbidity, as described (18). English Indices of Deprivation scores were calculated using postcodes from the current data provided by the UK’s Ministry of Housing, Communities and Local Government (2019) Report(19). Seven main types of deprivation are considered in the Index of Multiple Deprivation 2019 - income, employment, education, health, crime, access to housing and services, and living environment - and these are combined to form the overall measure of multiple deprivation.

**Table 1.** All patients with a confirmed positive severe acute respiratory syndrome coronavirus-2 (SARS-CoV-2) swab during study period. Legend. Data is number (percentage) unless otherwise stated. Ethnicity was self-reported or inferred (see Methods). Medical conditions were physician confirmed and checked against admission and linked primary care notes. English Indices of deprivation were calculated using postcode. Severity was determined by respiratory oxygen requirements, see methods. Subgroup data is provided for those ethnicities which represented more than 5% of the whole population. For English Indices of Deprivation, the quintiles were as follows; Quintile 1 = 33.5 – 78.1; Quintile 2 = 21.7 – 33.2; Quintile 3 = 14.4 – 21.5; Quintile 4 = 8.8 – 14.1; Quintile 5 = 1.4 – 8.6

|  | All patients | White Ethnicity | Asian Ethnicity | Black Ethnicity |
| --- | --- | --- | --- | --- |
| n | 2,217 | 1540 | 410 | 134 |
| Age in years: median (IQR) | 73 (58 - 84) | 77 (66 – 86) | 61 (45 - 73) | 62 (53 – 79) |
| Sex (n, %) |  |  |  |  |
| Female | 927 (41.8%) | 639 (41.5%) | 170 (41.5%) | 59 (44.0%) |
| Male | 1,290 (58.2%) | 901 (58.5%) | 240 (58.5%) | 75 (56.0%) |
| Self-reported ethnicity (n, %) |  |  |  |  |
| White | 1,540 (69.5%) | N/A | N/A | N/A |
| Mixed/ Multiple | 18 (0.8%) |  |  |  |
| South Asian/ South Asian British | 410 (18.5%) |  |  |  |
| Black/ African/ Caribbean/ Black British | 134 (6.0%) |  |  |  |
| Other ethnic group | 67 (3.0%) |  |  |  |
| Preferred not to say | 22 (1.0%) |  |  |  |
| Not known | 26 (1.2%) |  |  |  |
| Co-morbidity count (n, %) |  |  |  |  |
| None | 439 (19.8%) | 255 (16.6%) | 114 (27.8%) | 25 (18.7%) |
| 1 -2 | 888 (40.1%) | 620 (40.3%) | 155 (37.8%) | 61 (45.5%) |
| 3 or more | 890 (40.1%) | 665 (43.2%) | 141 (34.4%) | 48 (35.8%) |
| Morbidities (n, %) |  |  |  |  |
| Hypertension | 864 (39.0%) | 649 (42.1%) | 126 (30.7%) | 50 (37.3%) |
| Cerebrovascular Disease | 268 (12.1%) | 233 (15.1%) | 18 (4.4%) | 10 (7.5%) |
| Atrial fibrillation | 464 (20.9%) | 404 (26.2%) | 26 (6.3%) | 20 (14.9%) |
| Ischaemic heart disease, angina, myocardial infarct | 546 (24.6%) | 404 (26.2%) | 100 (24.4%) | 24 (17.9%) |
| Diabetes (type 1 and 2) | 752 (33.9%) | 434 (28.2%) | 197 (48.0%) | 72 (53.7%) |
| Asthma | 439 (19.8%) | 290 (18.8%) | 91 (22.2%) | 34 (25.4%) |
| COPD | 376 (17.0%) | 333 (21.6%) | 20 (4.9%) | 14 (10.5%) |
| Interstitial Lung Disease | 49 (2.2%) | 40 (2.6%) | 4 (1.0%) | 2 (1.5%) |
| Chronic Kidney Disease | 511 (23.0%) | 338 (21.9%) | 111 (27.1%) | 42 (31.3%) |
| Any active Malignancy | 152 (6.9%) | 124 (8.1%) | 12 (2.9%) | 7 (5.2%) |
| Dementia (all types) | 326 (14.7%) | 283 (18.4%) | 22 (5.4%) | 10 (7.5%) |
| Obesity | 267 (12.0%) | 174 (11.3%) | 60 (14.6%) | 12 (9.0%) |
| English Indices of Deprivation |  |  |  |  |
| 1 (most deprived) | 1,003 (45.2%) | 554 (36.0%) | 276 (67.3%) | 89 (66.4%) |
| 2 | 416 (18.8%) | 320 (20.8%) | 50 (12.2%) | 26 (19.4%) |
| 3 | 311 (14.1%) | 263 (17.1%) | 29 (7.1%) | 11 (8.2%) |
| 4 | 230 (10.3%) | 195 (12.7%) | 18 (4.4%) | 4 (3.0%) |
| 5 (least deprived) | 225 (10.1%) | 195 (12.7%) | 20 (4.9%) | 3 (2.2%) |
| Missing | 32 (1.4%) | 13 (0.8%) | 17 (4.2%) | 1 (0.8%) |
| Recovered and discharge (n, %) | 1,052 (47.5%) | 679 (44.1%) | 225 (54.9%) | 76 (56.7%) |
| Remain admitted at point of data census. (n, %) | 554 (25.0%) | 411 (26.7%) | 79 (19.3%) | 26 (19.4%) |
| Died (n, %) | 611 (27.5%) | 450 (29.2%) | 106 (25.9%) | 32 (23.9%) |
| Duration of symptoms prior to admission (in days)<br>n where data was available (% of 567) | 567 | 350 (61.7%) | 123 (21.7%) | 37 (6.5%) |
| **Median (IQR)** | 7 (3-10) | 6 (3 – 10) | 7 (4 – 10) | 7 (3.5 – 13.5) |
| **Severity of COVID on admission** |  |  |  |  |
| Documented (n,%) | 736 | 483 | 137 | 53 |
| Mild (n, % of those recorded) | 449 (61%) | 295 (61.1%) | 79(57.7%) | 36 (67.9%) |
| Moderate (n, % of those recorded) | 185 (25.1%) | 134 (27.7%) | 24(17.5%) | 10 (18.9%) |
| Severe (n, % of those recorded) | 102 (13.9%) | 54 (11.2%) | 34(24.8%) | 7 (13.2%) |
| Care escalation to ITU (n, %) | 269 (12.1%) | 133 (8.6%) | 86 (21.0%) | 21 (15.7%) |
| **Length of stay in full days (LOS): median (IQR)** |  |  |  |  |
| LOS for total population | 6 (2-12) | 6 (2-12) | 5 (2-9) | 5 (3-11) |
| LOS for patients discharged | 5 (2-11) | 6 (2-13) | 4.5 (2-8) | 5 (3-9) |
| LOS for patients who died | 6 (3-11) | 6 (3-12) | 5 (2-8) | 6 (3-9) |

Ethnicity was self-reported by the patient or their family members on admission to hospital. Where this data was missing, it was gathered from previous admissions and by reviewing primary and secondary medical records. If this was not available (as was the case in 91 patients), ethnicity was imputed from the modal ethnicity of patients with the same surname in the EHR database where possible, as previously described (20) but remained unavailable in 48 patients (see Table 1 for missing data). Ethnicity was grouped as per national guidelines(21).

### Severity of COVID-19 on admission

The physician determined severity of COVID-19 on first admission was categorised using a pragmatic and locally developed score which made use of baseline physiological assessments and oxygen requirements to identify those on admission to hospital who were in need of urgent critical care assessments for respiratory support and is as follows:

Patients were considered to have severe respiratory manifestations of COVID-19 infection; if COVID was suspected and the patient required inspired oxygen > 50% to maintain targeted oxygen saturations (>93% except in the presence of type 2 respiratory failure where the target saturations were 88% - 92%) with respiratory pathology thought driven by COVID-19 illness.

If not severe, patients were considered to have moderate severity respiratory manifestations of COVID-19 infection if COVID was suspected and the patient required inspired oxygen of > 4L/min or inspired oxygen > 28% to maintain target oxygen saturations.

Patients were considered to have mild severity respiratory manifestations of COVID-19 infection if the patient had respiratory symptoms but did not meet the severe or moderate criteria as described above.

Baseline physiological assessments to determine severity of COVID-19 were considered to be those taken within 24 hours either side of the SARS-CoV-2 swab collection time, of which the earliest available measurement was used. Since not all patients were admitted within 24 hours of their SARS-CoV-2 swab test, and since these assessments are only routinely recorded in the EHR system for QEHB patients, baseline severity scores were only available for a subset of the patients (736/2217).

To determine if disease severity on admission reflected duration of illness, medical clerking notes were reviewed to determine the duration of symptoms prior to admission. This was available in only a subset of patients (567/2217).

### Outcomes

The primary outcome was death while in hospital or post discharge until 12th May 2020. For those patients discharged from hospital, primary care records were checked and any patients admitted to hospital with COVID-19 and discharged who had died in the community within the censor period were noted. Those with an on-going admission were censored on the study end date.

### Statistics

#### Baseline Characteristics

Baseline characteristics for the total population and ethnic communities are presented as mean (standard deviation) or median (interquartile range) for continuous variables and as frequency (percentage) for categorical variables. Ethnic groups were compared by age, sex, co-morbidity and severity on presentation.

#### Standardised Admission Rate and Standardised Mortality Rate by ethnicity

Ethnicity data for the Birmingham and Solihull area from the 2011 census were used to estimate expected numbers of admissions and deaths for each ethnic category conditional on the observed numbers and sex-specific age distributions of COVID-19 admissions and deaths in UHB. The ratios of observed to expected numbers were calculated to provide standardised admission ratios (SAR) and standardised mortality ratios (SMR) for each ethnic category and 95%, 99% and 99.9% confidence intervals were obtained using the mid-P exact test(22).

#### Predictors of mortality

The overall effects of age, sex, ethnicity, co-morbidities and admission severity on mortality were tested by univariable analysis. The effect of ethnicity on mortality was then considered adjusting for age, sex, comorbidity counts and deprivation in a multivariable analysis. Cox model was used to derive adjusted Hazard Ratios (aHR) for mortality, defined as death from any cause after COVID-19. Survival time was calculated as the time between the collection of a sample on clinical suspicion, to the date of death or study end date and used for Kaplan-Meier estimates. Multiple parametrisations were tested for patient age, including linear fit, square-root transformation, categorical groupings, and natural cubic splines. Categorical variables were fitted for sex, ethnicity, deprivation score quintiles and number of clinically assessed co-morbidities. The proportional hazards assumption was tested through correlation of the scaled Schoenfeld residuals with survival time, with hypothesis tests for independence (23). Models were then tested using Propensity Score Matching (24) where age, sex, social deprivation index and co-morbidities were matched and ethnicity was treated as a dichotomous variable. Models were fitted in R version 3.6.3 using the survival package and twang package in “r” (www.r-project.org)(25). All p values are reported exactly and no corrections were made for multiple comparisons unless stated.

#### Patient and public involvement

302 patients and public members were consulted as to the use of health data to improve the care for people with acute, unplanned illness. A group of patients recovering from COVID-19 specifically were asked and supported the use of routinely collected health data to investigate the relationship between poor outcomes and ethnicity. A working group of staff and patients from Black and Asian Minority ethnic groups discussed the results and how they should be disseminated.

## Results

2,217 consecutive patients admitted to UHB with a swab proven diagnosis of COVID-19 were analysed. A modified CONSORT diagram (Figure S1 of the online supplement) and a summary of the demographic and clinical characteristics on admission (Table 1) are shown. Most patients (n=2132) had a Birmingham postcode. The majority of patients were male (1290/2217; 58.2%) and White (1540/2217; 69.5%). High levels of co-morbidity were identified across all ethnic groups (with 40.1% of patients having three or more co-morbidities).

Of note, up to 50% had missing morbidity data and 30% had missing ethnicity data using the secondary care records for the COVID-related admission alone, but >96% of ethnicity data and all available morbidity data was resolved through reviewing primary and secondary care records.

### Comparison of baseline characteristics between ethnicities

South Asian patients, when compared to White patients, were younger (median age 61 years vs. 77 years; p<0.001), had a higher prevalence of diabetes mellitus (48.1% vs 28.2%, p<0.001) but lower prevalence of dementia (5.4% vs 18.9%), COPD (4.9% vs 21.6%), atrial fibrillation (6.3% vs 26.2%) and cerebrovascular disease (4.4% vs 15.1%) (p<0.0001 for all). Similar to the South Asian patients, Black ethnic population were younger (median age 62 years), had a higher prevalence of diabetes mellitus (53.7%) but lower prevalence of other key co-morbidities than the White population. Two thirds of the South Asian and Black ethnicity came from the most deprived quintile of deprivation compared to 36% from the White patients.

Across the course of data collection, the proportion of patients presenting from different ethnic groups was relatively stable (Figure S2 of the online supplement), suggesting no differential transmission related to ethnic group within the location and timeframe studied.

### Standardised Admission Ratio in different ethnicities

Age and sex Standardised Admission Ratio (95% Cl) for South Asian females was 74% higher (SAR, 1.7: 95%CI 1.5-2.0) and for South Asian males 63% higher (SAR, 1.6: 95%CI 1.4-1.9) than the standard population. (See Table S1 of the online supplement). In contrast White patients were less likely to be admitted in comparison to the standard population (White females; 0.8 (0.8 - 0.9) and White males; 0.9 (0.8 - 0.9)). Admission rates were similar to expected rates of the standard population for Black ethnicity.

### Severity of COVID19 disease at presentation in different ethnicities

In 736 patients admitted directly to the QEHB site, the severity of COVID-19 recorded on admission identified 185 (25.1%) classified as moderate and 102 (13.9%) classified as severe. In this dataset a higher proportion of South Asians than Whites were assessed to have severe disease on presentation (34/137 (24.8%) vs 54/483 (11.2%) p <0.0001). See table S2 of the online supplement. A higher proportion of South Asians than Whites were admitted to ICU (86/410 (21.0%) vs 133/1540 (8.6%) p <0.001). There were no differences in the duration of symptoms prior to admission by ethnic group ((Median) South Asian = 7 days, White = 6 days, Black =7 days, p=0.40).

### Characteristics of patients who died

On 12^th^ May 2020, 769/2217 (34.6%) patients had died in hospital or following discharge. These patients were older, more likely to be male, White, have multiple co-morbidities and in the 736 patients admitted directly to the QEHB site, in whom data was available, more likely to have moderate or severe disease on admission, in comparison to all other groups of patients (table 2).

**Table 2:** A comparison of demographics in patients who met the primary endpoint of death up to and including on 12 ^th^ May 2020. Legend. Data is number (percentage) unless otherwise stated. Medical conditions were self-reported and checked against admission and linked primary care notes. Groups are compared using Chi-squared analysis, except for pregnancy where Fisher’s exact test was used and age distribution, where Kruskal Wallis was used. Of note, p values compared all patients currently alive (in patients or discharged) versus those who had died. * = Increased deaths with 3 or more (p<0.0001) co-morbidities on post hoc analysis. ** = increased deaths in moderate and severe cases vs. mild cases on post hoc analysis (p < 0.0001)for comparisons with patients discharged alive and currently alive. For English Indices of Deprivation, the quintiles were as follows; Quintile 1 = 33.5–78.1; Quintile 2 = 21.7 – 33.2; Quintile 3 = 14.4 – 21.5; Quintile 4 = 8.8 – 14.1; Quintile 5 = 1.4–8.6

|  | **Patients currently alive (admitted or discharged)** | **Patients discharged and alive** | **Patients still admitted/ receiving in-patient care and alive** | **Patients who died** | **P value comparing all alive patients and patients who died** |
| --- | --- | --- | --- | --- | --- |
| n | 1,448 | 1,372 | 76 | 769 |  |
| Age (IQR and range) | 68 (52-80) | 68 (52 – 80) | 66 (54 – 76) | 80 (71 - 887) | $p<0.001$ |
| Sex (n, % female) | 637 (44.0%) | 616 (44.9%) | 21 (27.6%) | 290 (37.7%) | $p=0.004$ |
| **Self-reported ethnicity (n, %)** | | | | | $p<0.001$ |
| White | 958 (66.2%) | 906 (66.0%) | 52 (68.4%) | 582 (75.7%) |  |
| Mixed/ Multiple | 10 (0.7%) | 10 (0.7%) | 0 (0.0%) | 8 (1.0%) |  |
| South Asian/ South Asian British | 290 (20.0%) | 275 (20.0%) | 15 (19.7%) | 120 (15.6%) |  |
| Black/ African/ Caribbean/ Black British | 94 (6.5%) | 92 (6.7%) | 2 (2.6%) | 40 (5.2%) |  |
| Other ethnic group | 60 (4.1%) | 53 (3.9%) | 7 (9.2%) | 7 (0.9%) |  |
| Preferred not to say | 16 (1.1%) | 16 (1.2%) | 0 (0.0%) | 6 (0.8%) |  |
| Unknown | 20 (1.4%) | 20 (1.5%) | 0 (0.0%) | 6 (0.8%) |  |
| **Co-morbidity count** | | | | | $p<0.0001$ |
| None | 377 (26.0%) | 362 (26.4%) | 15 (19.7%) | 62 (8.1%) |  |
| 1 or 2 | 593 (41.0%) | 555 (40.5%) | 38 (50.0%) | 295 (38.4%) |  |
| 3 or more | 478 (33.0%) | 455 (33.2%) | 23 (30.3%) | 412 (53.6%)* |  |
| **Morbidities** |  |  |  |  |  |
| Hypertension | 490 (33.8%) | 457 (33.3%) | 33 (43.4%) | 374 (48.6%) | $p<0.001$ |
| Cerebrovascular Disease | 128 (8.8%) | 125 (9.1%) | 3 (3.9%) | 140 (18.2%) | $p<0.001$ |
| Atrial fibrillation | 236 (16.3%) | 228 (16.6%) | 8 (10.5%) | 228 (29.6%) | $p<0.001$ |
| Ischaemic heart disease, angina, myocardial infarct | 286 (19.8%) | 276 (20.1%) | 10 (13.2%) | 260 (33.8%) | $p<0.001$ |
| Diabetes (type 1 and 2) | 459 (31.7%) | 431 (31.4%) | 28 (36.8%) | 293 (38.1%) | $p=0.002$ |
| Asthma | 296 (20.4%) | 280 (20.4%) | 16 (21.1%) | 143 (18.6%) | $p=0.299$ |
| COPD | 202 (14.0%) | 194 (14.1%) | 8 (10.5%) | 174 (22.6%) | $p<0.001$ |
| Interstitial Lung Disease | 24 (1.7%) | 23 (1.7%) | 1 (1.3%) | 25 (3.3%) | $p=0.015$ |
| Chronic Kidney Disease | 267 (18.4%) | 253 (18.4%) | 14 (18.4%) | 244 (31.7%) | $p<0.001$ |
| Any active Malignancy | 87 (6.0%) | 84 (6.1%) | 3 (3.9%) | 65 (8.5%) | $p=0.030$ |
| Dementia (all types) | 131 (9.0%) | 129 (9.4%) | 2 (2.6%) | 195 (25.4%) | $p<0.001$ |
| Obesity | 176 (12.2%) | 163 (11.9%) | 13 (17.1%) | 91 (11.8%) | $p=0.825$ |
| **English Indices of deprivation** |  |  |  |  |  |
| 1 | 667 (46.1%) | 633 (46.1%) | 34 (44.7%) | 336 (43.7%) | $p=0.848$ |
| 2 | 271 (18.7%) | 253 (18.4%) | 18 (23.7%) | 145 (18.9%) |  |
| 3 | 200 (13.8%) | 191 (13.9%) | 9 (11.8%) | 111 (14.4%) |  |
| 4 | 150 (10.4%) | 146 (10.6%) | 4 (5.3%) | 80 (10.4%) |  |
| 5 | 139 (9.6%) | 128 (9.3%) | 11 (14.5%) | 86 (11.2%) |  |
| Missing | 21 (1.5%) | 21 (1.5%) | 0 (0.0%) | 11 (1.4%) |  |
| **Duration of symptoms prior to admission (in days) n where data was available Median (IQR)** | 245 (16.9%)<br>9 (2-16) | 223 (16.3%)<br>9 (3-16) | 22 (28.9%)<br>7 (5-12) | 110 (14.3%)<br>7 (3-15) |  |

**Table 2: A comparison of demographics in patients who met the primary endpoint of death up to and including on 12<sup>th</sup> May 2020.**
| Severity of COVID on admission (n,%) | n = 489 | n = 453 | n = 36 | n = 247 | p<0.001 |
| --- | --- | --- | --- | --- | --- |
| Mild | 348 (71.2%) | 338 (74.6%) | 10 (27.8%) | 101 (40.8%) |  |
| Moderate | 91 (18.6%) | 81 (17.8%) | 10 (27.8%) | 94 (38.1%) |  |
| Severe | 50 (10.2%) | 34 (7.5%) | 16 (44.4%) | 52 (21.1%) ** |  |

In those in whom data was available, people that died had a shorter duration of symptoms prior to admission compared to those who were still alive (median 7 days (3 - 36) vs. 9 (5 - 36) days). In all in whom data was available, the duration of symptoms prior to admission did not related to disease severity on admission (p=0.46). The relationships between survival from diagnosis and age, gender and number of co-morbidities are further illustrated in Figure 1.

**Figure 1:**
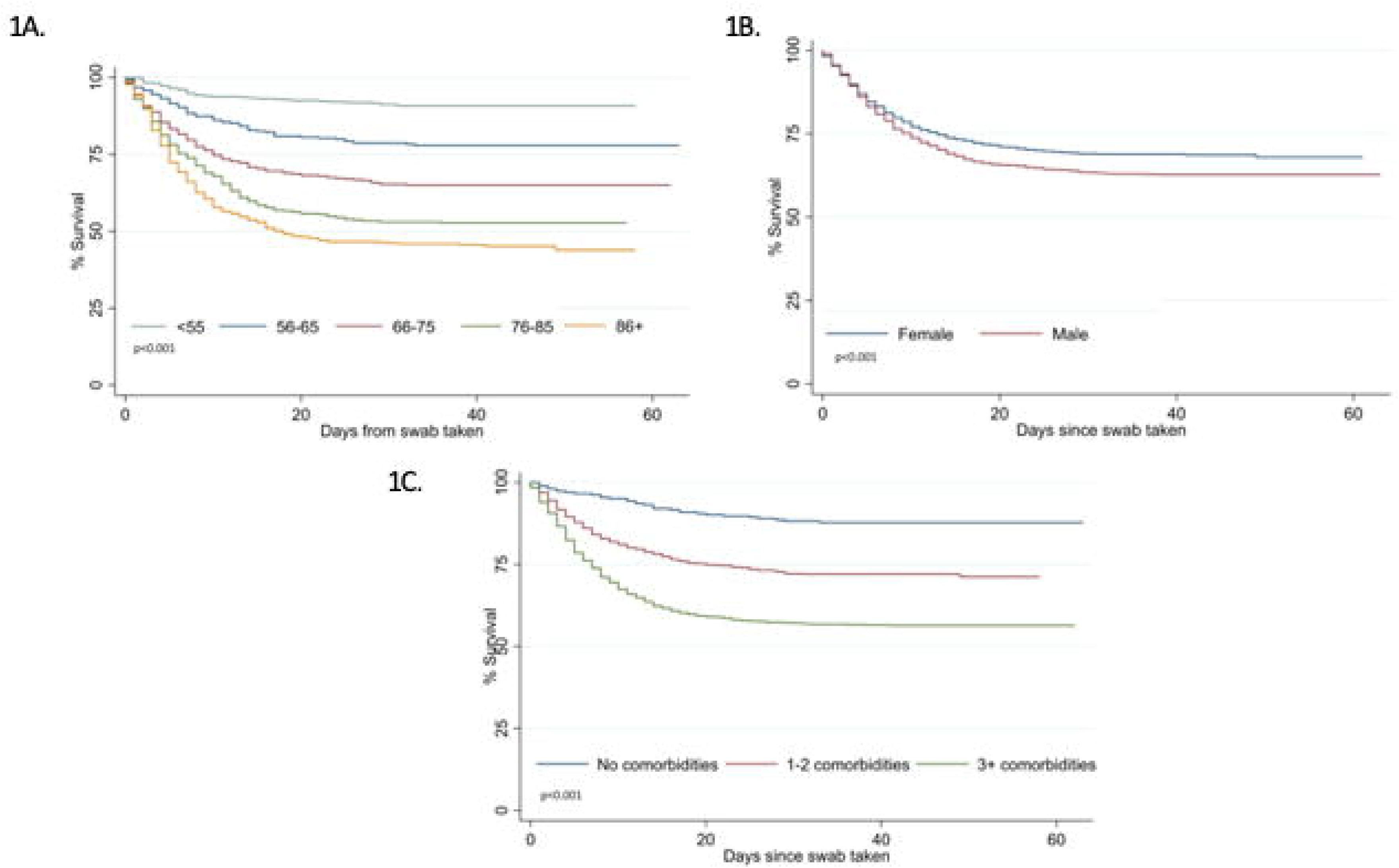
Kaplan-Meier estimates of survival for COVID positive patients. **Legend**. Data compares survival status of patients by age (figure 1A, p <0.001); sex (figure IB, p <0.001) and simple co-morbidity counts as listed in table 1 (figure 1C: p <0.001). Comparison using the log-rank test

### Ethnic group outcome analysis

#### Standardised Mortality Ratio between different ethnic groups

In comparison to the expected number of deaths based on Birmingham and Solihull 2011 census data age and sex structure, there were significantly more South Asian women and men who died with a positive COVID-19 swab than would be expected: Standardised Mortality Ratios (SMR) (95% Cl): Asian females; 1.9 (1.4-2.6): Asian males; 1.7 (1.4 - 2.1). In contrast fewer White women and men died than would be expected: SMR (95% Cl): White females; 0.9 (0.8 - 1.0): White males; 0.9 (0.8 - 1.0). (See online Supplementary table S1). For those of Black ethnicity, death rates were not different to the expected rates in the standard population.

#### Survival curves for mortality in different ethnicities

An age adjusted Kaplan-Meier showed that although there were no differences in age -adjusted survival in White and Black ethnicities, patients from Asian ethnic groups were less likely to survive (See Figure 2).

**Figure 2:**
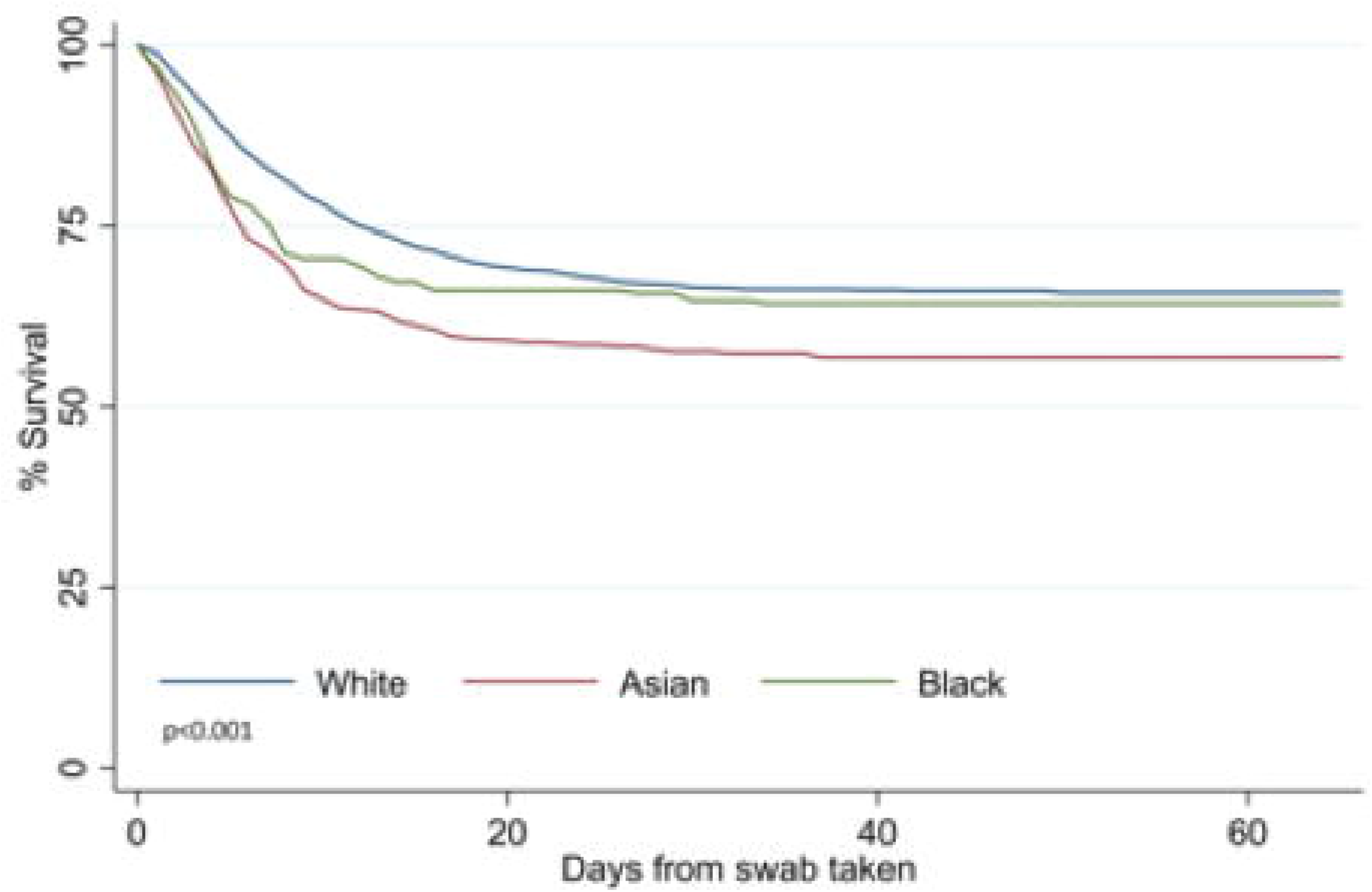
Age adjusted Kaplan-Meier estimates of survival for different ethnic groups of COVID positive patients. Legend. Data compares age-adjusted survival status of patients by ethnicity (p <0.001 using the age ranges as listed in Figure 1A). Comparison using the log-rank test

#### Multivariable analysis

In a multivariable Cox regression model adjusted for age, sex, comorbidity counts and deprivation, South Asian ethnicity (aHR 1.4, 95%CI: 1.2-1.8) was associated with a significantly higher risk of death. Within the limits of the power of the study there was absence of a significant difference in survival for Black ethnicity compared to the White population (aHR 1.1, 95%CI: 0.8-1.5). In addition to this we found age z-score (aHR 2.4: 95%CI 1.8-3.2) and co-morbidities (1-2 comorbidity, aHR 1.7: 95%CI 1.3-2.2; and 3 or more comorbidities, aHR 2.3: 95%CI 1.7-3.0) as significant predictors. We also found a significant effect of sex (aHR for male sex, 1.3, 95% Cl: 1.1-1.5), with the interaction between age and sex suggesting an amplified risk in males with increasing age (Table 3). The main effects of age and multiple co-morbidities were modified by an interaction (aHR for age interaction with 1-2 comorbidities, 0.9, 95%CI: 0.7-1.2; for age interaction with 3+ comorbidities, 0.7, 95% Cl: 0.5-0.9), that attenuated the relative impact of increasing comorbidity at advanced age. Of note, the HR of 1.4 (1.2-1.8), did not change (1.4 (1.2-1.8)), if the interactions were removed from the model.

**Table 3.**
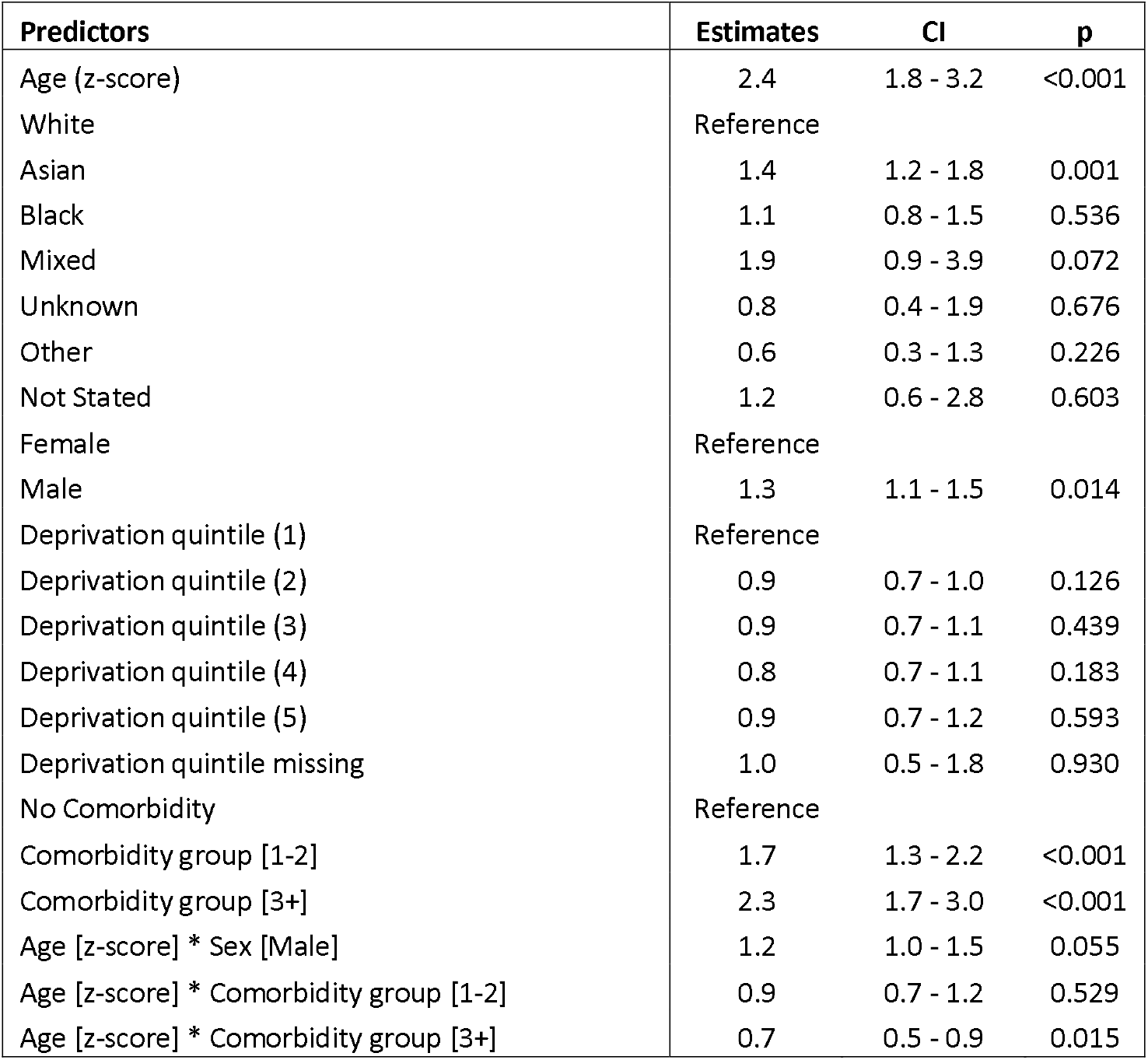
Adjusted Hazard Ratios of risk factors for mortality. **Legend**. Multivariable Cox regression model including age (z-score), ethnicity, sex and comorbidity count as covariates. Adjusted Hazard Ratios along with their confidence intervals are presented.

| Predictors | Estimates | CI | p |
| --- | --- | --- | --- |
| Age (z-score) | 2.4 | 1.8 - 3.2 | <0.001 |
| White | Reference |  |  |
| Asian | 1.4 | 1.2 - 1.8 | 0.001 |
| Black | 1.1 | 0.8 - 1.5 | 0.536 |
| Mixed | 1.9 | 0.9 - 3.9 | 0.072 |
| Unknown | 0.8 | 0.4 - 1.9 | 0.676 |
| Other | 0.6 | 0.3 - 1.3 | 0.226 |
| Not Stated | 1.2 | 0.6 - 2.8 | 0.603 |
| Female | Reference |  |  |
| Male | 1.3 | 1.1 - 1.5 | 0.014 |
| Deprivation quintile (1) | Reference |  |  |
| Deprivation quintile (2) | 0.9 | 0.7 - 1.0 | 0.126 |
| Deprivation quintile (3) | 0.9 | 0.7 - 1.1 | 0.439 |
| Deprivation quintile (4) | 0.8 | 0.7 - 1.1 | 0.183 |
| Deprivation quintile (5) | 0.9 | 0.7 - 1.2 | 0.593 |
| Deprivation quintile missing | 1.0 | 0.5 - 1.8 | 0.930 |
| No Comorbidity | Reference |  |  |
| Comorbidity group [1-2] | 1.7 | 1.3 - 2.2 | <0.001 |
| Comorbidity group [3+] | 2.3 | 1.7 - 3.0 | <0.001 |
| Age [z-score] * Sex [Male] | 1.2 | 1.0 - 1.5 | 0.055 |
| Age [z-score] * Comorbidity group [1-2] | 0.9 | 0.7 - 1.2 | 0.529 |
| Age [z-score] * Comorbidity group [3+] | 0.7 | 0.5 - 0.9 | 0.015 |

To further test the assumption that South Asian ethnicity was associated with worse outcomes even when co-morbidities, age and sex were considered, propensity score matching was conducted, matching for the same factors in the Cox regression but categorising ethnicity into South Asian or not. With propensity score matching the Hazard ratio for risk of death was 1.3 (1.0-1.6) for South Asian patients compared with non-South Asian patients. When co-morbidities were added independently (rather than as a count) using propensity score matching, South Asian ethnicity was still associated with a significantly higher risk of death (HR 1.4 (95% CI 1.1-1.7)).

## Discussion

This is the first study to specifically describe the impact of South Asian ethnicity on the outcome of COVID-19 infection using highly characterised and accurate primary and secondary data from patients admitted to hospital in the UK. South Asians were significantly younger and twice likely to have diabetes than White patients and when accounted for the age structure of the local population, had a high admission and death rate. South Asians were also more likely to present with severe symptoms but with no difference in the duration of symptoms and more likely to be admitted to ITU. Importantly, after adjusting for age, deprivation, and multiple co-morbidities the effect of South Asian ethnicity on mortality was 42% higher.

Our study is in line with the two population based study (10,11), where South Asians were found to be at increased risk of death with similar effect size with the study(ll) that considered similar covariates for adjusting (aHR 1.4 (1.2-1.8) vs 1.6 (1.4-1.8). However, the findings of the two population-based studies could reflect infection rates rather than case fatality rates and therefore differentiating this is important from a public health and research perspective. If increased deaths in South Asian patients reflected high infection rates, then our focus should be on looking at barriers and emphasising the need for adherence to current social distancing guidelines. If this reflects an increased susceptibility to poorer outcomes from SARS-CoV-2 infection, we need to urgently understand the reasons for severity of infection and mitigate risk or develop targeted treatments. While no firm conclusion can be drawn from the current dataset, our study potentially supports the latter, with South Asian patients more likely to be admitted, more likely to present with severe symptoms and have an increased risk of mortality. This supports the call by the government and research community for urgent research on the reasons underpinning these observations. Our study was not sufficiently powered to report on other ethnic groups, particularly those of Black ethnicity, and therefore the findings among these ethnic groups should be interpreted cautiously.

The excess age-adjusted mortality in COVID-19 is not solely attributable to a range of cardiovascular and metabolic risk factors that are over-represented in this ethnic group. In sensitivity analysis the Hazard Ratio for South Asian ethnicity using both Cox regression and Propensity score matching was stable when ‘number of co-morbidities’ was exchanged for the presence of specific co-morbidities including diabetes mellitus and hypertension. To place this in context, the effect of South Asian on mortality is significantly less than the effect of one or more co-morbidities (present in 80.2% of all admitted patients) and approximates to the effect of ageing ten years in the White population. In this study we did not observe an independent signal related to higher deprivation levels(26) and poor outcomes in contrast to the population based study(ll), suggesting deprivation is likely to be related to high infection rates and thereby high mortality rather than high rates of severe infection leading to increased mortality.

It is notable that in the sub-analysis of patients admitted to QEHB, where we were able to immediately integrate a COVID-19 specific assessment into our electronic healthcare record, South Asian patients appear to present with more severe disease but there was no difference in the duration of symptoms prior to admission, suggesting disease severity was not caused simply by delayed presentation to medical services or differences in health service utilisation (although this cannot be fully excluded, given the unknown burden of COVID-19 in the community). Indeed, when comparing those still alive and those who had died at the end of the study, the patients who had died had a shorter history of symptoms prior to admission, suggesting a different disease course.

A significantly higher rate of admission to ITU in the South Asian ethnic group could relate to this more severe disease at presentation. It may also relate to patient level differences in joint decision-making regarding ITU treatment, in patients who have higher levels of specific co-morbidities such as dementia and COPD, groups that are significantly over-represented in the White ethnic group which was also significantly older.

The limitations in our overall analysis need to be considered, specifically that 5% of patients remain in hospital at the time of the data lock and more patients have been admitted, so our findings will evolve. Since the proportion of patients presenting from different ethnic groups was stable across the course of data collection, any consequence for our main conclusion on the mortality risk in South Asians admitted to hospital is likely to be small. Data on the admission severity scoring did not include all patients, which is a limitation. This limitation reflects the real-world response within a global pandemic which includes designing a score to inform care escalation decisions and updating the UHB electronic health record to capture this information during the first wave of patient admissions. However, these data suggest further exploration of severity of disease at presentation is warranted.

It is also important to acknowledge that standardised admission and mortality ratios from Birmingham and Solihull use the most recent census data, but that these are from 2011. Estimates of the contemporary age structure do not however suggest a need to significantly qualify these findings. The UK has not undertaken widespread screening or diagnosis of patients in the community, we are therefore unable to comment upon the natural history of COVID-19 prior to admission to secondary care, irrespective of ethnic group. This testing regime is likely to evolve with the development of capacity and methodology and will provide a more complete picture of COVID-19. A description of disease in the community will help build a clearer understanding of the apparent excess mortality following admission, for which there remain a number of possible explanations. A limitation of this study (and, arguably, of any observational study) is that it cannot exclude the possibility that another, unmeasured variable could account for the ethnicity effect described here. The assessment of co-morbidities does not reflect the degree of severity of the condition, nor disease treatment or control, and the assessment of social deprivation might impact on chronic disorders. There is also the possibility that differences in work or home living arrangements might impact on potential transmission and this may be different across ethnic communities. However, a real strength has been the ability to study a highly curated and complete data set, without the inherent issues of significant under-coding seen with morbidity and ethnicity data when using a secondary care dataset.

The biological basis of any difference in outcome can only be speculated upon at present. There are reported differences in outcomes for non-white ethnic groups from ARDS even after adjusting for sex, age, disease severity, type of hospital, and median household income(27, 28). The worst clinical manifestations of COVID-19 appear to be associated with a cytokine storm syndrome. Here a hypercytokinaemia is seen (4), with predictors of mortality reflecting a virally induced inflammatory state which can be assessed using a scoring system including validated clinical laboratory tests (29). Candidate genes associated with ARDS have been identified in bioinformatic analyses with a strong predominance of inflammatory pathways, including reactive oxygen species, innate immunity-related inflammation, and endothelial vascular signalling pathways (30). Ethnicity may influence cytokine gene polymorphisms and inflammatory profiles following specific challenges (31) with some ethnic groups more prone to a heightened inflammatory response. Of note, socio-economic factors might also impact on inflammatory pathways and gene expression(32). These factors remain poorly understood, were a priority for our patient and public involvement group who were consulted for this study, and there is an urgent need to understand the genomic and associated phenomic and socio-economic characteristics of patients who are susceptible or resistant to the severe manifestations of COVID-19 to understand this further.

Although our study includes only one NHS Foundation Trust, it covers an ethnically diverse contiguous population of 1.3 million people for which it is the sole provider of adult acute secondary care across 4 hospital sites. This provides for continuity of data, clinical protocols and access to therapy. The immediate availability of access to an electronic representation of a primary care record to support the care of admitted patients, also supports the integrity of data collection, the quality of which might otherwise be more limited (33).

Our findings, which describe and quantify the risk of COVID19 in the South Asian population are relevant to national policy and to understanding the underlying biological mechanisms in ‘at risk’ populations. Future studies will extend our observations and explore underlying epidemiology and biological mechanisms; to improve interventions based in the community, the emergency department, ward and ITU. Perhaps most importantly our findings inform the UK’s national discussion on at ‘at risk’ groups and the ensuing fear arising from uncertainty.

## Data Sharing Agreement

To facilitate knowledge in this area, the anonymised participant data and a data dictionary defining each field will be available to others through application to PIONEER via the corresponding author.

## Author contribution

E Sapey designed the study, collated data, performed some analysis, wrote the manuscript. S Gallier collated data, performed analysis, wrote the manuscript. C Mainey performed statistical analysis. P Nightingale performed statistical analysis and helped write the manuscript. D McNulty performed statistical analysis. H Crothers performed statistical analysis. F Evison performed statistical analysis. K Reeves performed statistical analysis. D Pagano assisted with the design of the study and manuscript preparation. P. Diggle assisted with analysis. K Nirantharakumar and AK Denniston assisted with analysis and manuscript writing. S Ball designed the study, oversaw data collection, helped write the manuscript.

All authors amended the manuscript and approved the final version. The data was curated and analysed on behalf of all clinicians at UHB.

## Licence for Publication

The Corresponding Author has the right to grant on behalf of all authors and does grant on behalf of all authors, an exclusive licence (or non exclusive for government employees) on a worldwide basis to the BMJ Publishing Group Ltd to permit this article (if accepted) to be published in BMJ Open Respiratory Research and any other BMJPGL products and sublicences such use and exploit all subsidiary rights, as set out in our licence (http://group.bmj.com/products/journals/instructions-for-authors/licence-forms).

## Data Availability

To facilitate knowledge in this area, the anonymised participant data and a data dictionary defining each field will be available to others through application to PIONEER via the corresponding author or HDR-UK Innovation Gateway. The data will be available upon request and following approval of a process to ensure ethical data governance and through a data access agreement. Please contact the corresponding author for details.

## Acknowledgements

This work was supported by PIONEER, the Health Data Research UK (HDR-UK) Health Data Research Hub in acute care. HDR-UK is an initiative funded by UK Research and Innovation, Department of Health and Social Care (England) and the devolved administrations, and leading medical research charities. This work uses data provided by patients and collected by the NHS as part of their care and support. We would like to acknowledge the contribution of all staff, key workers, patients and the community who have supported our hospitals and the wider NHS at this time

The manuscript was prepared and submitted on behalf of all of the staff and students at UHB NHS Foundation Trust

With particular thanks to

Medical Students: Lylah Irshad, Maxim Harris, Theodore Nabav’,

Health Informatics: A Kolesnyk, M Ahmed A Liaqat

Tanya Pankhurst, Jamie Coleman, Chirag Dave, Khaled ElFandi, Rifat Rashid, Paul Cockwell

## Research in context

### What is the key question

Are some ethnic groups more at risk of worse outcomes from COVID-19 infection, including death?

### What is the bottom line

There were more admissions from South Asian patients to our hospital than would be expected based on our local population. These patients were admitted with a worse severity of COVID19 related respiratory compromise without a significant delay in presentation and experience a higher level of mortality even when differences in age, sex, deprivation and key comorbidities were taken into account.

### Why read on

South Asian ethnicity may form another ‘at risk’ population from COVID-19 and further studies are needed to identify any treatable factors to improve outcomes as well as to refine our understanding and communication around non-modifiable risk factors.

## References

1. Phelan AL, Katz R, Gostin LO. The Novel Coronavirus Originating in Wuhan, China: Challenges for Global Health Governance. JAMA. 2020;323(8):709–10.

2. World Health Organisation. Novel Coronavirus(2019-nCoV) Situation Report - 11. https://www.whoint/docs/default-source/coronaviruse/situation-reports/20200131-sitrep-ll-ncovpdf?sfvrsn=de7c0f74. 2020;DOA 6.4.2020.

3. Chen T, Wu D, Chen H, Yan W, Yang D, Chen G, et al. Clinical characteristics of 113 deceased patients with coronavirus disease 2019: retrospective study. BMJ. 2020;368:m1091.

4. Huang Y, Yang R, Xu Y, Gong P. Clinical characteristics of 36 non-survivors with COVID-19 in Wuhan, China. medRxiv. 2020:2020.02.27.20029009.

5. Onder G, Rezza G, Brusaferro S. Case-Fatality Rate and Characteristics of Patients Dying in Relation to COVID-19 in Italy. JAMA. 2020.

6. Richardson S, Hirsch JS, Narasimhan M, Crawford JM, McGinn T, Davidson KW, et al. Presenting Characteristics, Comorbidities, and Outcomes Among 5700 Patients Hospitalized With COVID-19 in the New York City Area. JAMA. 2020.

7. Centers of Disease Control and Prevention. Non Hispanic Black People Disproportionately Affected by COVID-19 Hospitalizations in CDC Data. https://www.cdcgov/coronavirus/2019-ncov/cases-updates/. 2020; DOA 11.4.2020.

8. Intensive Care National Audit and Reseach Centre. ICNARC report on COVID-19 in critical care 10th April. https://www.icnarcorg/Our-Audit/Audits/Cmp/Reports. 2020;DOA 12.4.2020.

9. Centre. ICNAaR. ICNARC report on COVID-19 in critical care 17th April. https://www.icnarcorg/Our-Audit/Audits/Cmp/Reports. 2020; DOA 21.4.2020.

10. Office for National Statistics. Coronavirus (COVID-19) related deaths by ethnic group, England and Wales: 2 March 2020 to 10 April 2020. https://www.onsgovuk/peoplepopulationandcommunitv/birthsdeathsandmarriages/deaths/articles/coronavirusrelateddeathsbvethnicgroupenglandandwales/2march2020tol0april2020. 2020;DOA 12.5.2020.

11. OpenSafely Collaborative;. OpenSAFELY: factors associated with COVID-19-related hospital death in the linked electronic health records of 17 million adult NHS patients. https://opensafelvorg/outputs/2020/05/covid-risk-factors/. 2020;DOA 12.5.2020.

12. Office for National Statistics. CT0430_2011 Census - Sex by age by country of birth by ethnic group - Birmingham LA and Solihull LA combined. https://www.onsgovuk/peoplepopulationandcommunitv/populationandmigration/populationestimates/adhocs/004295ct04302011censussexbvagebvcountrvofbirthbvethnicgroupbirminghamlaandsolihulllacombined. 2011;DOA 10.4.2020.

13. Carver K, Jones N. Compartive accuracy of oropharyngeal and nasopharyngeal swabs for diagnosis of COVID-19. Oxford COVID-19 Evidence Service Team Centre for Evidence-Based Medicine, Nuffield Department of Primary Care Health Sciences University of Oxford. 2020;https://www.cebm.net/covid-19/comparative-accuracv-of-oropharvngeal-and-nasopharvngeal-swabs-for-diagnosis-of-covid-19/.

14. NHS England. Guidance and standard operating procedure COVID-19 virus testing in NHS laboratories. NHS England and Improvements. 2020;https://www.england.nhs.uk/coronavirus/wp-content/uploads/sites/52/2020/03/guidance-and-sop-covid-19-virus-testing-in-nhs-laboratories-vl.pdf.

15. NHS Digital. The NHS Digital SNOMED CT Browser. https://termbrowsernhsuk/? 2020;DOA 10.4.2020.

16. Organisation. WH. ICD-10 online versions. https://www.whoint/classifications/icd/icdonlineversions/en/. 2019;DOA 3.3.2020.

17. Caramelo F, Ferreira N, Oliveiros B. Estimation of risk factors for COVID-19 mortality - preliminary results. medRxiv. 2020:2020.02.24.20027268.

18. Guan W-j, Liang W-h, Zhao Y, Liang H-r, Chen Z-s, Li Y-m, et al. Comorbidity and its impact on 1590 patients with Covid-19 in China: A Nationwide Analysis. European Respiratory Journal. 2020:2000547.

19. Ministry of Housing Calg. The English Indices of Deprivation 2019 Technical report. https://deraioeacuk/34259/l/loD2019TechnicalReportpdf. 2019;DOA 10.4.2020.

20. Gagne F. Discriminant Function Analysis. Biochemical Ecotoxicology: Principles and methods. 2014;Academic Press.

21. Office for National Statistics. Ethnic group, national identity and religion. Measuing equality: a guide for the collection and classification of ethnic group, national identity and religion data in the UK. https://www.onsgovuk/methodology/classificationsandstandards/measuringequalitv/ethnicgroupnationalidentitvandreligion. 2016;DOA 20th March 2020.

22. Dean AG, Sullivan KM, Soe MM. OpenEpi: Open spurce epidemiological statistics for public health. Updated 2013/04/06. www.OpenEpicom. 2013;DOA 2020/04/19.

23. Schoenfeld D. Chi-squared goodness-of-fit tests for the proportional hazards regression model. Biometrika. 1980;67(1):145–53.

24. Austin PC. An Introduction to Propensity Score Methods for Reducing the Effects of Confounding in Observational Studies. Multivariate Behavioral Research. 2011;46(3):399–424.

25. Therneau TM. Package Survival. https://cranuibno/web/packages/survival/survivalpdf. 2020;DOA 19.4.2020.

26. Khunti K, Singh AK, Pareek M, Hanif W. Is ethnicity linked to incidence or outcomes of covid-19? BMJ. 2020;369:m1548.

27. Erickson SE, Shlipak MG, Martin GS, Wheeler AP, Ancukiewicz M, Matthay MA, et al. Racial and ethnic disparities in mortality from acute lung injury. Critical care medicine. 2009;37(1):1–6.

28. Bime C, Poongkunran C, Borgstrom M, Natt B, Desai H, Parthasarathy S, et al. Racial Differences in Mortality from Severe Acute Respiratory Failure in the United States, 2008-2012. Annals of the American Thoracic Society. 2016;13(12):2184–9.

29. Mehta P, McAuley DF, Brown M, Sanchez E, Tattersall RS, Manson JJ. COVID-19: consider cytokine storm syndromes and immunosuppression. The Lancet. 2020;395(10229): 1033–4.

30. Lynn H, Sun X, Casanova N, Gonzales-Garay M, Bime C, Garcia JGN. Genomic and Genetic Approaches to Deciphering Acute Respiratory Distress Syndrome Risk and Mortality. Antioxid Redox Signal. 2019;31(1557-7716 (Electronic)): 1027–52.

31. Hoffmann S, Stanley E, Cox E, DiMercurio B, Koziol D, Harlan D, et al. Ethnicity Greatly Influences Cytokine Gene Polymorphism Distribution. American journal of transplantation: official journal of the American Society of Transplantation and the American Society of Transplant Surgeons. 2002;2:560–7.

32. Thames AD, Irwin MR, Breen EC, Cole SW. Experienced discrimination and racial differences in leukocyte gene expression. Psychoneuroendocrinology. 2019;106(1873-3360 (Electronic)):277–83.

33. Iqbal G, Gumber A, Johnson MRD, Szczepura A, Wilson S, Dunn JA. Improving ethnicity data collection for health statistics in the UK. Diversity in Health and Care. 2009;6: https://diversitvhealthcare.imedpub.com/improving-ethnicitv-data-collection-for-health-statistics-in-the-uk.php?aid=2045.

